# Identifying the critical time window for the association of the early-life gut microbiome and metabolome with childhood neurodevelopment

**DOI:** 10.1101/2021.12.30.21268329

**Authors:** Zheng Sun, Kathleen Lee-Sarwar, Rachel S. Kelly, Jessica A. Lasky-Su, Augusto A. Litonjua, Scott T. Weiss, Yang-Yu Liu

## Abstract

It has been widely recognized that a critical time window for neurodevelopment occurs in early life, and that the host’s gut microbiome plays an important role in neurodevelopment. While murine models have demonstrated that the maternal gut microbiome also influences offspring brain development, for humans it is still unclear if the critical time window for the association between the gut microbiome and neurodevelopment is prenatal, postnatal or both. Here we leverage a large-scale human study and compare the associations between the gut microbiota and metabolites from mothers and their children with the children’s neurodevelopment. We show, for the first time, that the maternal gut microbiome is more relevant than the children’s gut microbiome to the children’s neurodevelopment in the first year of life. Interestingly, the roles of the same taxa with respect to neurodevelopment can be opposite at the two stages of fetal neurodevelopment. These findings shed light on potential therapeutic interventions to prevent neurodevelopmental disorders.

## INTRODUCTION

Mounting evidence suggests that the gut microbiome influences not only the host’s health status^1^ but also brain function and host behavior^2,3^. In particular, neurodevelopment has been shown to be associated with gut microbial composition in animal models^4-8^. For example, compared with normal mice, germ-free mice differ in social behavior, stress response, cognition, and other functions, providing solid evidence for the role of the microbiome in neurodevelopment^9^. Some of these effects can be mitigated by exposure to microbes throughout adolescence, while others persist despite microbial colonization^10^, suggesting that early life is a sensitive time period for the effects of microbial exposure on neurodevelopment^11^. Since a human child’s brain grows to 80–90% of its adult volume by two years of age^12^, it is crucial to identify the critical time window for the association between the gut microbiome and neurodevelopment in the early years.

In addition to the relationship between the infant gut microbiome and neurodevelopment, previous studies in mice^13-15^ and humans^16^ suggest that the maternal microbiome also influences brain development in offspring. However, it is still unclear if the impact of the maternal microbiome is limited to events during pregnancy or occurs postnatally as a result of vertical transmission^17-19^. A recent mouse study revealed that the maternal gut microbiome promotes fetal thalamocortical axonogenesis by signaling neurons in the developing brain of the offspring with microbially modulated metabolites^20^. However, the relative importance of the association between prenatal (or postnatal) gut microbiome and neurodevelopment in humans remains to be determined.

Although mouse studies are important in providing evidence supporting the concept that the gut microbiome is involved in neurodevelopment^4-11,13-15,20,21^, there are significant limitations to human translation of these findings^22^. Microbiome data in human populations are clearly warranted, especially with longitudinal data allowing exploration of the vertical transmission from mother to child. In this study, we leveraged microbiome and metabolome (microbiome for short) data from the Vitamin D Antenatal Asthma Reduction Trial (VDAART^23^), which assessed the effect of vitamin D supplementation in pregnancy on development of asthma-related phenotypes in offspring. Specifically, we assessed associations between the maternal (and child) gut microbiome on children’s neurodevelopment from year one to year three of life as measured by the Ages and Stages Questionnaire, third edition (ASQ-3, or ASQ for short hereafter)^24^ (see **Methods**). More specifically, we examined the predictive power of (*i*) the maternal gut microbiome, (*ii*) the infant gut microbiome (3 to 6 months) and (*iii*) the gut microbiome of children at year one and year three on outcomes of children’s neurodevelopment (**Fig.1**). The comparison of relevance (as measured by their predictive power for the ASQ) of maternal, infant and child gut microbiome allowed us to identify the earliest, most primary association between the gut microbiome and the neurodevelopment in humans. Interestingly, we found that the maternal gut microbiome is more relevant to their children’s first-year neurodevelopment than the infant (months 3-6) gut microbiome or the gut microbiome at year one, while the predictive power of maternal and infant gut microbiome gradually diminishes for neurodevelopment at year two. Neither the maternal nor children’s microbiome has any predictive power for the children’s neurodevelopment at year three. Our results underscore the importance of maternal gut microbiome in offspring neurodevelopment, which may guide us in designing potential interventions.

**Figure 1.**
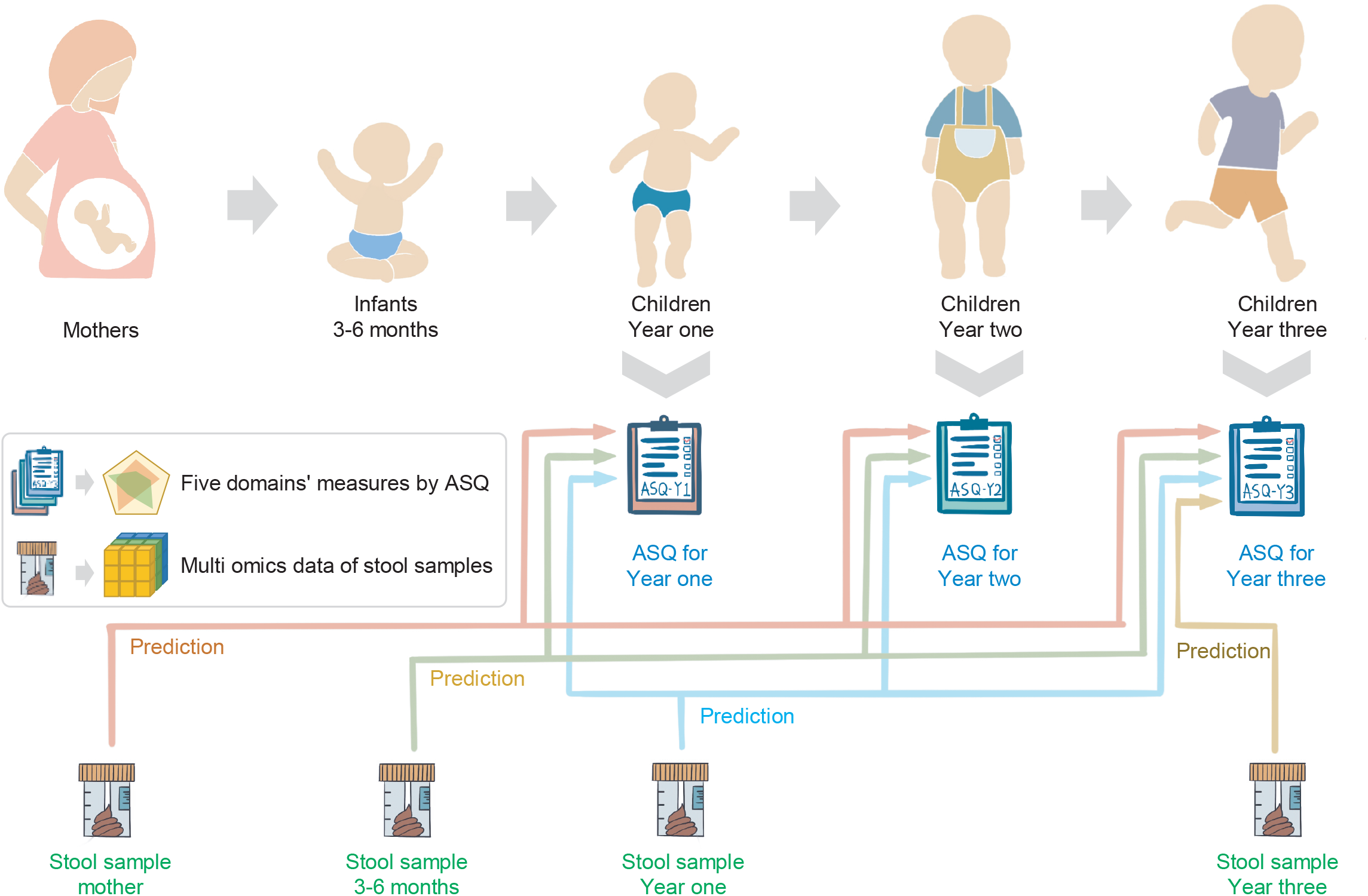
Analysis design. Conceptual diagram illustrating the analysis design and collection of stool samples and ASQ questionnaires. The gut microbial compositions of mothers, infants (months 3-6), and children (year one and year three) were separately used to predict neurodevelopmental outcomes as measured by ASQ at different ages (ASQ-Y1, ASQ-Y2, and ASQ-Y3) to quantify the associations and relevance between the gut microbiome and neurodevelopment.

## RESULTS

### Participants and data collection

In the VDAART cohort, 876 pregnant women were recruited from three sites across the United States between 2009 and 2015 and their offspring continue to be followed (**Methods**). In this study, 116 mother-child pairs were selected for their relatively complete multi-omics datasets and ASQ questionaries (**Methods, Fig.S1**): (*i*) The stool metagenomic sequencing data are accessible for 116 mothers, 85 infants at months 3 to 6, 105 children at year one, and 80 children at year three; (*ii*) The stool short-chain fatty acid (SCFA) measurements are accessible for 113 mothers, 73 infants at months 3 to 6, 68 children at year one, and 65 children at year three; (*iii*) The stool metabolomic profiles are available for 111 mothers, 114 infants at months 3 to 6, 75 children at year one, and 70 children at year three. As for the ASQ measures of the children, completed ASQ data are available for 99 children at year one (ASQ-Y1), 102 children at year two (ASQ-Y2), and 110 children at year three (ASQ-Y3).

The maternal participants (n=116) are ethnically diverse (22% Hispanic or Latino and 78% not Hispanic or Latino) and racially diverse (38% white, 49% black, and 13% other, **Table 1**). They also exhibit a representative range of family incomes (36% less than 30k/year, 30% 30k-100k/year, and 9% more than 100k/year) and education levels (62% attended at least some college). In terms of health status, 35% of mothers had asthma, 28% had eczema, and 56% had a fever during pregnancy. Most of their children (65%) were delivered vaginally and more than half were breastfed (52%) in the first six months of life. In addition, 48% of mothers were randomized to receive high-dose vitamin D during pregnancy. All the above clinical covariates that might be associated with the gut microbiota or neurocognitive development were considered as potential confounders in the downstream statistical analysis.

**Table 1.** Characteristics of mother participants in this study. Data are given as number (and percentage) of individuals. Besides mothers’ characteristics, their children’s information such as gender, ethnicity and race are also included. Not all metagenomic and metabolomic data are available for all 116 mother-child pairs, please see **Fig.S1** for detailed information.

| Characteristic and mothers # (n=116) |  |  | Characteristic and mothers # (n=116) |  |  |
| --- | --- | --- | --- | --- | --- |
| Recruitment site |  |  | Eczema |  |  |
| Boston | 52 | (45%) | Yes | 32 | (28%) |
| St Louis | 49 | (42%) | No | 84 | (72%) |
| San Diego | 15 | (13%) | Asthma |  |  |
| Maternal education level |  |  | Yes | 41 | (35%) |
| College graduate or Graduate school | 37 | (32%) | No | 75 | (65%) |
| Some college | 35 | (30%) | High fever |  |  |
| High school or technical school | 24 | (21%) | Yes | 65 | (56%) |
| Less than high school | 20 | (17%) | No | 51 | (44%) |
| Family income |  |  | Treatment |  |  |
| > 100K/year | 10 | (9%) | Vitamin D | 61 | (53%) |
| 30K-100K/year | 35 | (30%) | Blank | 55 | (47%) |
| < 30K/year | 42 | (36%) | Delivery mode |  |  |
| Unknown | 29 | (25%) | Cesarean Section | 41 | (35%) |
| Marital status |  |  | Vaginal birth | 75 | (65%) |
| Single | 64 | (55%) | Child Gender |  |  |
| Married | 47 | (41%) | Female | 52 | (45%) |
| Divorced | 5 | (4%) | Male | 64 | (55%) |
| Maternal ethnicity |  |  | Child Ethnicity |  |  |
| Hispanic or Latino | 26 | (22%) | Hispanic or Latino | 30 | (26%) |
| Not Hispanic or Latino | 90 | (78%) | Not Hispanic or Latino | 86 | (74%) |
| Maternal race |  |  | Child Race |  |  |
| Black or African American | 57 | (49%) | Black or African American | 66 | (57%) |
| White | 44 | (38%) | White | 36 | (31%) |
| Other | 15 | (13%) | Other | 14 | (12%) |
| Gestation days |  |  | Breast feed |  |  |
| 30-33 weeks | 57 | (49%) | Yes | 60 | (52%) |
| 34-37 weeks | 59 | (51%) | No | 56 | (48%) |

### Assessment of children’s neurodevelopment by ASQ and profiling of the gut microbiome at different stages

To evaluate children’s neurodevelopment, ASQ measures at year one, year two, and year three were analyzed. Scores within the domains of communication skills (COM), gross motor skills (GM), fine motor skills (FM), problem-solving ability (ProS), and personal social skills (PerS) were summed to give an overall continuous measurement for each of the five domains (see **Methods**). We found that the ASQ scores over the three years measured were unstable and unpredictable whether separated into individual domains or combined in a single summary score (**Fig.2**), which is in line with previous studies^25^. For example, subjects with lower scores at year one (ASQ-Y1) may have higher scores at year two (ASQ-Y2) and may again have lower scores at year three (ASQ-Y3). Thus, it is inappropriate to categorize the children based on the irregular change of the three years’ ASQ measures. Hence, the association between the gut microbiota and ASQ measures were separately analyzed year by year in this study.

**Figure 2.**
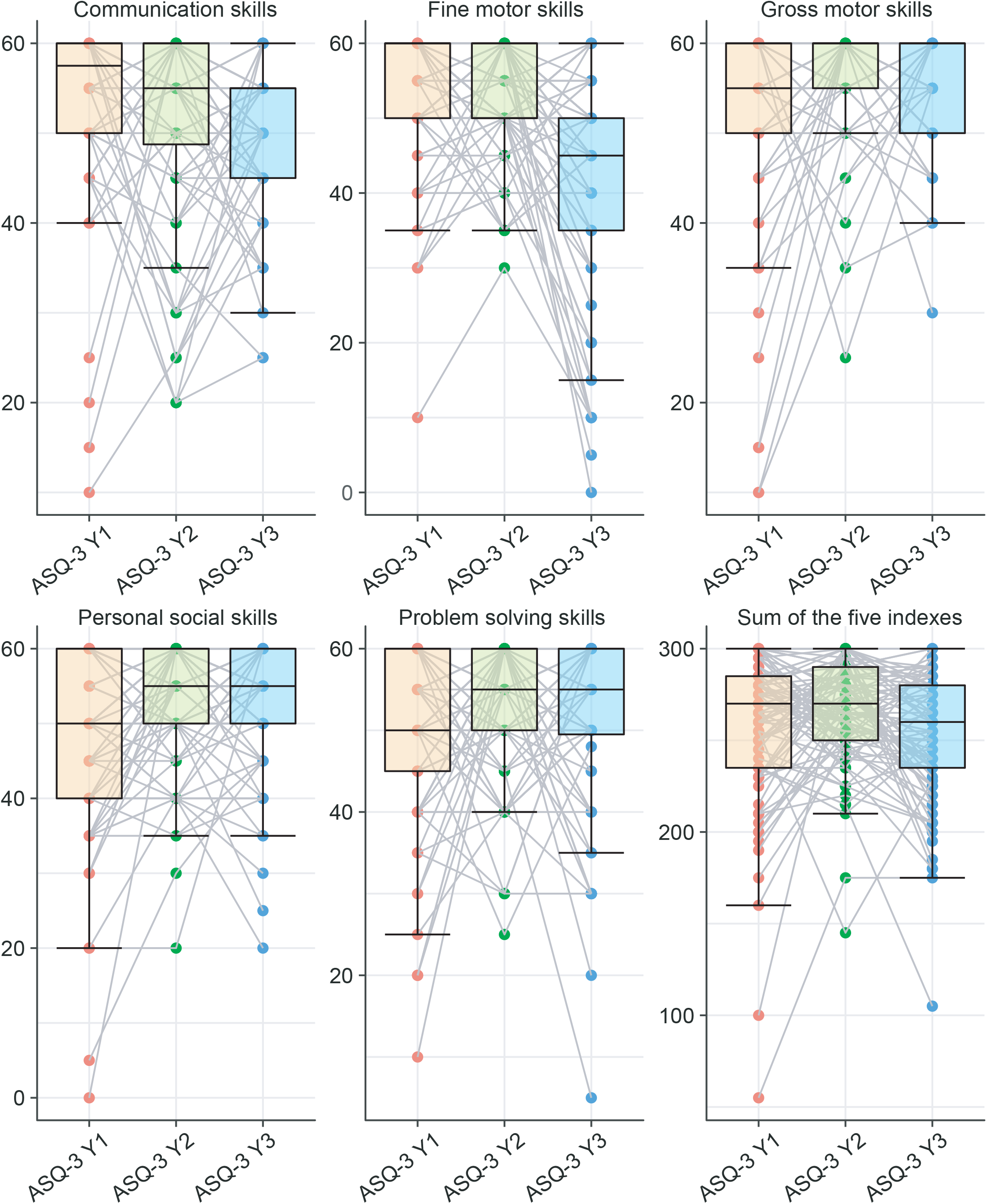
The change of the five domains in ASQ form year one to year three. The boxplots illustrate the comparison of the five ASQ measures and their sum score across the first three years of life, with grey lines connecting the same individual. ASQ-Y1 (orange boxes) refers to the ASQ at year one, ASQ-Y2 (green boxes) refers to the ASQ at year two, and ASQ-Y3 (blue boxes) refers to the ASQ at year three.

We analyzed the maternal, infant (months 3-6) and children’s year one and year three gut microbial compositions. We found that the taxonomic diversity increased along with the children’s age. In particular, we identified 3,770 ASVs (Amplicon Sequence Variants) in the maternal gut microbiome, 1,440 ASVs in the infant gut microbiome, 2,376 ASVs in child gut microbiome at year one, and 2,360 ASVs in child gut microbiome at year three (please see **Fig.S2** for overall picture (PCoA) of the microbiome and metabolome data at different stages).

### The prenatal period may be the critical time window for the association between the gut microbiome and neurodevelopment

To further identify associations of the maternal gut microbiome, the infant gut microbiome (months 3-6), and the child gut microbiome (year one and year three) with neurodevelopment (ASQ-Y1, ASQ-Y2, and ASQ-Y3), we performed multinomial regression using reference frames^26^ to explicitly address the compositionality issue in microbiome data analysis. Specifically, we compared null models with series of multinomial regression models built for predicting the neurodevelopment of different ages (ASQ-Y1, ASQ-Y2, and ASQ-Y3) based on the gut microbiota and metabolites of different stages (mother, infant, child year one, and child year three). The Q^2^ score (adapted from the partial least squares literature) allows us to see how strongly the ASQ measures are associated with the gut microbiome at different stages, as compared to random chance (**Methods**). Higher Q^2^ (positive) indicates a higher predictive accuracy (with values of 1 corresponding to 100% accuracy) on cross-validation and suggests a stronger relationship between the gut microbiome and neurodevelopment, while negative Q^2^ indicates poor predictive accuracy and irrelevance.

When using the maternal gut microbiome to predict the ASQ measures (**Fig.3a, Fig.S3a**), we found that the best performance of predicting the five domains of ASQ-Y1 by the maternal gut microbiota is at the class-level (with average Q^2^ ∼ 0.21 compared to 0.145, 0.127, 0.099, 0.018, and 0.018 at the phylum, order, family, genus and ASV level, respectively). Moreover, both the maternal gut microbiota and metabolites are significantly associated with the five domains in children’s ASQ-Y1, with Q^2^ ranging from 0.190 to 0.233 for microbiota at the class level and Q^2^ ranging from 0.108 to 0.157 for metabolites at the pathway level (**Methods**). Of note, the performance of predicting ASQ-Y2 (or ASQ-Y3) measures by the maternal gut microbiome is much lower than that of predicting ASQ-Y1 measures (with Q^2^ values less than 0.052 and 0.039 for ASQ-Y2 and ASQ-Y3).

**Figure 3.**
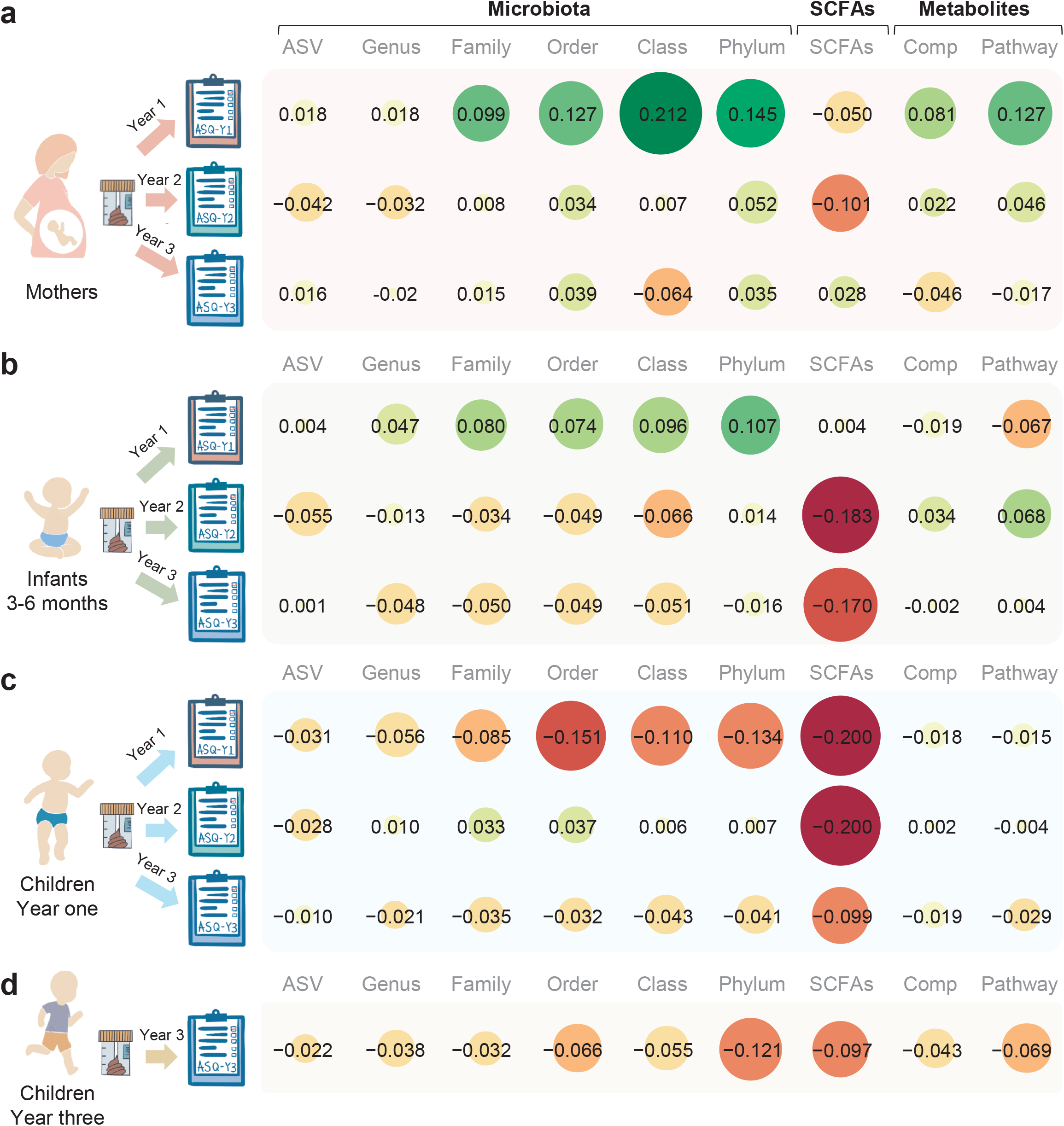
Average predictive power (Q^2^) of the gut microbiome for the five domains in ASQ. (**a**) Predictive power of the maternal gut microbiome for child neurodevelopment. (**b**) Predictive power of the infant gut microbiome in early life (months 3-6) for child neurodevelopment. (**c**) Predictive power of the child gut microbiome at year one for child neurodevelopment. (**d**) Predictive power of the child gut microbiome at year three for child neurodevelopment at the same time point. Red pies indicate negative Q^2^ and green pies indicate positive Q^2^. The number in each pie chart is the average Q^2^ of communication skills, fine motor skills, gross motor skills, personal social skills, and problem-solving skills. The higher Q^2^ (positive) indicates a higher predictive accuracy for ASQ, while negative Q^2^ indicates poor predictive accuracy or overfitting of the multinomial regression models for ASQ measures. The value less than -0.2 will be trimmed to -0.2 for visualization, and comp is short for compound (metabolite).

When using the infant gut microbiome to predict the ASQ measures (**Fig.3b, Fig.S3b**), we found that the infant gut microbiota is only relevant to ASQ-Y1 (Q^2^ ranges from 0.076 to 0.128 at the class level) while the metabolites are only relevant to ASQ-Y2 (Q^2^ ranges from 0.036 to 0.094 at the pathway level), and the SCFAs showed limited relevance to FM, GM and ProS of ASQ-Y1. Of note, prediction of ASQ-Y1 measures using the infant gut microbiota (with average Q^2^ ∼ 0.096) is much weaker than that of using the maternal gut microbiota (with average Q^2^ ∼ 0.212). The performance of predicting ASQ-Y3 measures using the infant gut microbiome is very low (with Q^2^ values less than 0.004).

When using the child gut microbiome (year one and year three, **Fig.3c,3d, Fig.S3c**,**3d**) to predict the ASQ measures, we found that the year one child gut microbiome (including SCFAs) is not relevant for predicting ASQ-Y1 but slightly relevant for predicting ASQ-Y2 (Q^2^ ranges from to 0.011 to 0.065 for the five domains in ASQ-Y2 at the order level). We also found that year-three child gut microbiome (including SCFAs) cannot predict ASQ-Y3 measures (with Q^2^ values less than -0.02).

A high consistency finding by metagenomic and metabolomic data indicate that the maternal gut microbiome is much more relevant for predicting the offspring year-one neurodevelopment (assessed by ASQ-Y1 measures) than the infant (months 3-6) and the child gut microbiome (year one). Also, the predictive power of maternal and infant gut microbiome gradually diminishes with respect to neurodevelopment at year two (assessed by ASQ-Y2) and neither the maternal nor child microbiome has any predictive power for the neurodevelopment at year three (assessed by ASQ-Y3).

### The different roles of the maternal and infant gut microbiota in neurodevelopment

To further explore whether the predictive power of the infant gut microbiome for neurodevelopment is a consequence of vertical transmission from mothers to their children, we analyzed the similarity of the gut microbiota between paired mothers and children (**Fig.4a**). We found that the similarity of the infant gut microbiota with their mother increased along with age. For example, the average Bray-Curtis dissimilarity of the gut microbiota between infant and their mother is 0.904, while for the child gut microbiota at year one and year three, at which points the microbiome was poorly predictive of ASQ-Y1 (or ASQ-Y3, **Fig.3c**), the number is 0.806 and 0.678, respectively. This suggests that the relevance between the infant gut microbiota and ASQ measures is not associated with the similarity with the maternal gut microbiota. Thus, the roles of the maternal and infant gut microbiomes in neurodevelopment are potentially different.

**Figure 4.**
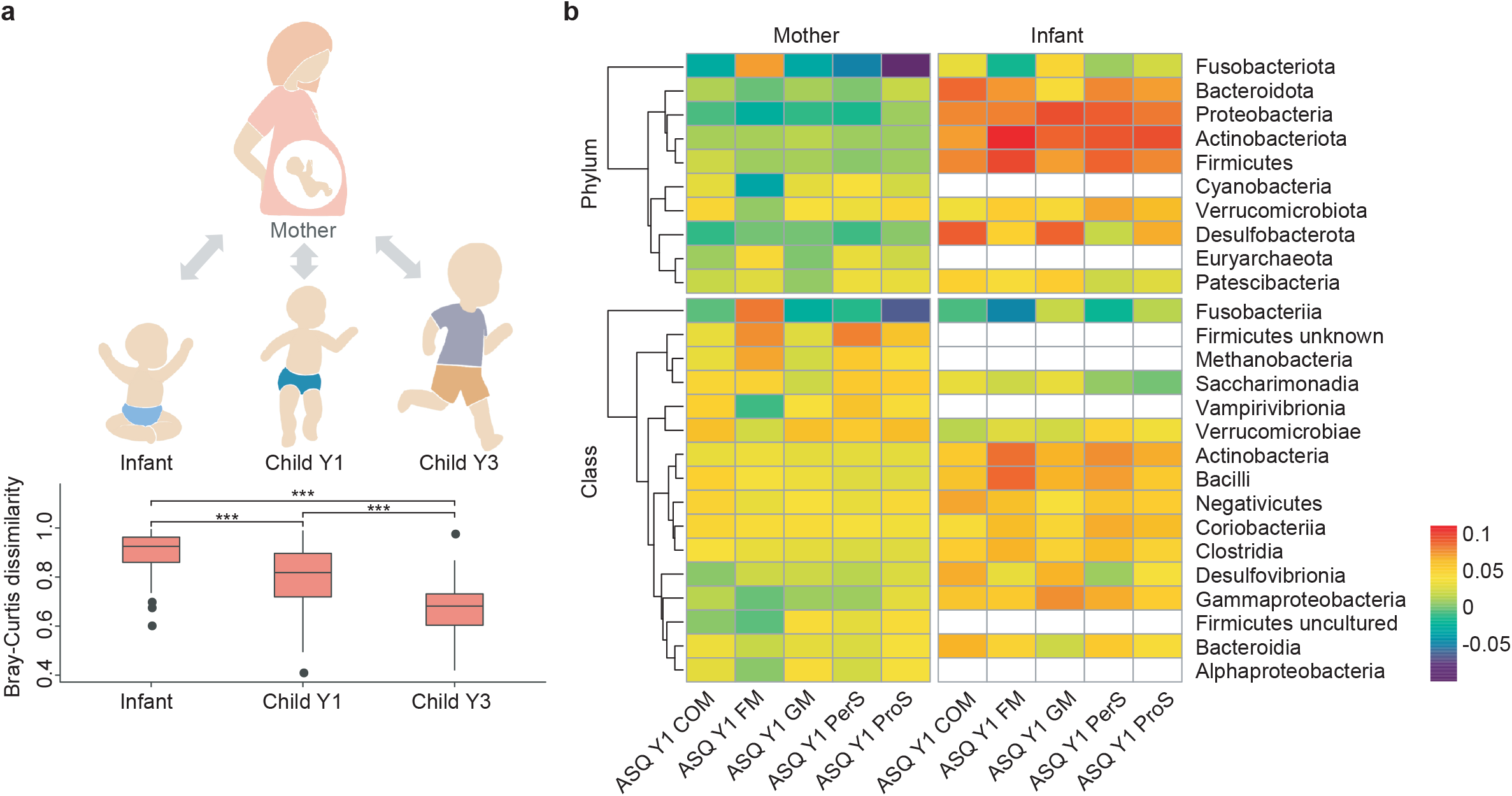
Different roles of maternal and infant gut microbiota in neurodevelopment. (**a**) Comparison of BC dissimilarity between paired maternal and infant gut microbiota. Red boxes refer to the BC dissimilarity between paired mothers and their children (n=85, 105, 80 for infant, Child Y1 and Child Y3). (**b**) The heatmap shows the association ranks of classes from maternal, infant and children gut microbiota against ASQ scores.

To further understand the role of the gut microbiota in neurodevelopment, we compared the differentials (the term “differential” refers to the logarithm of the fold change in abundance of a taxa with respect to ASQ measures^26^, see **Methods**) with respect to the five domains in ASQ-Y1 from the maternal and infant gut microbiota. Specifically, since the microbial taxa at the class and phylum level from these two stages are most predictive of ASQ-Y1, we compared the ranks of differential classes and phyla in multinomial regression models, which give information on the relative associations of taxonomic classes and phyla with ASQ-Y1 (**Methods**). Although the differential taxa in the maternal and infant gut with respect to ASQ-Y1 exhibit high overlap (e.g., there are 11/16 and 8/10 ASQ-Y1 related classes and phyla shared between the two stages, **Fig.4b**), our results demonstrated that the same taxa in the maternal and infant gut microbiota do not necessarily play the same role in neurodevelopment. For example, the class Fusobacteriia is more associated with high fine motor skills in ASQ-Y1 (rank = 0.084) in the maternal gut microbiota, but become more associated with low fine motor skills (rank = -0.047) in the infant gut microbiota (the relative abundance of Fusobacteriia is 0.02% and 0.03% in the maternal and infant gut microbiota, **Fig.4b**). Moreover, in the maternal gut microbiota, the phylum Fusobacteriota is mostly associated with high scores in the five domains in ASQ-Y1 (rank for COM, FM, GM, PerS, and ProS are -0.030, 0.073, -0.031, -0.049, and -0.099 respectively) while Fusobacteriota exhibits opposite associations (0.033, -0.017, 0.048, 0.009, and 0.024) in the infant gut microbiota for neurodevelopment.

## DISCUSSION

Mounting evidence linking offspring neurodevelopment with the maternal gut microbiota comes from mouse studies^4-11,13-15,20,21^. To date, only one human study has examined the role of the maternal gut microbiota in offspring neurodevelopment^16^. More studies are certainly needed to address foundational questions about the maternal gut microbiota and children’s neurodevelopment. To our knowledge, this is the first human study with comprehensive multi omics data on this topic. In this work, we leveraged a human study to compare the ability of the maternal and child gut microbiomes to predict child neurodevelopment. Interestingly, the maternal gut microbiome exhibited stronger associations with child neurodevelopment at year one than the infant (months 3-6) or the first-year child gut microbiomes. Further analysis illustrated that similar class-level taxa were associated with neurodevelopment among both the maternal and infant gut microbiota, but those taxa may play different roles during pregnancy and in infancy.

A previous study found structural differences in the maternal gut microbiota between the mothers of children with behavioral problems (n=20) in comparison to mothers of normative behavior children (n=195)^16^, consistent with our finding of a significant effect of the maternal gut microbiota on ASQ measures (including problem solving skills and personal social skills). In our study, instead of using the Childhood Behavior Checklist at age two (which may lead to unbalanced case-control sample size and subsequent problematic differential abundance analysis), we measured neurodevelopment by ASQ in the first three years of life (ASQ-Y1, ASQ-Y2, and ASQ-Y3) and then quantitatively associated its five domain scores with maternal and child gut microbiome features at different time points (months 3-6, year one, and year three), allowing us to explore associations of the gut microbiome with neurodevelopment more comprehensively.

As both 16S rRNA and whole metagenome sequencing data in microbiome studies are inherently compositional, differential abundance analysis is challenging. Indeed, analyzing relative abundance data with inappropriate statistical methods can lead to 100% false discovery rates^26-28^. Comparing ratios of taxa as relative differentials (logarithm of the fold change in abundance of taxa between two conditions) can circumvent this issue. Compared with previous studies about the association between maternal gut microbiota and child neurodevelopment, we applied reference frames to address the compositionality inherent in microbiome data^26^. Specifically, we estimated the relative differentials directly using multinomial regression^29-31^, then compared our model with a null model to quantify the relevance between features (the profiling results of the gut microbiota) and factors (ASQ measures), and the coefficients from multinomial regression analysis were ranked to determine which taxa varied the most along with ASQ measures.

In previous studies, *Fusobacterium*, Fusobacteriaceae, Fusobacteriales, Fusobacteria, and Fusobacteriota have been reported to be associated with brain and nervous system disorders like major depressive disorder^32^ and multiple sclerosis^33^. In addition, the relative abundance of Fusobacteria was found to be different in rats who are vulnerable to early life stress^34^. However, although validation and mechanistic studies are needed to confirm our findings, we report for the first time a potential two-sided role of Fusobacteria and Fusobacteriota in neurodevelopment with divergent associations with ASQ-Y1 when measured in pregnant mothers compared to during infancy. As for the rest of the shared differential classes between the maternal and infant gut microbiota with respect to the five domains in ASQ-Y1, many of them, including Desulfovibrionia, Gammaproteobacteria, Negativicutes, Bacteriodia, Verrucomicrobiae, Clostridia, Bacilli, Actinobacteria, and Coribacteria, have been related to neurodevelopment in previous studies^35-41^, which supports the accuracy of our overall findings.

There are several limitations in this study. For example, the sample size is limited, and more time points evaluated during pregnancy are required to narrow down the critical time window for offspring neurodevelopment. Moreover, the relative abundance of Fusobacteriia is extremely low in this study, further validation of its two-sided role by qPCR or digital PCR in larger population is highly warranted. In addition, future research utilizing multi-omics data (e.g., whole metagenomic sequencing which would be useful to evaluate the role of strain-level vertical transmission) and systematic experiments are further warranted to deepen our understanding of the mechanism of prenatal and postnatal influence on neurodevelopment by the gut microbiome.

## METHODS

### VDAART study design and sample selection

Subjects were offspring of participants in VDAART, a multi-site randomized, double-blind, placebo-controlled trial of Vitamin D supplementation during pregnancy for prevention of asthma and other allergic disease in offspring conducted in the United States (NCT00920621)^23^. The study protocol was approved by the institutional review boards at each participating institution and at Brigham and Women’s Hospital. All participants provided written informed consent. Pregnant women (n=876) were recruited during the first trimester of pregnancy from three sites across the United States: Boston Medical Center, Washington University in St. Louis, and Kaiser Permanente Southern California Region in San Diego. After delivery, 806 children were followed using an over-the-phone quarterly health questionnaire and annual in-person visits. Stool samples were collected from mothers at the third trimester, and from their children at months 3-6, age one and age three years.

Stool was not collected if the mother or child had used antibiotics in the past 7 days. Then metagenomic sequencing, metabolomic profiling, and measurement of short chain fatty acids were conducted on the collected stool samples as detailed below (see **Fig.1** for detailed sample size of each omics datasets).

### Ages and Stages Questionnaire

At the end of the first, second and third year, study researchers administered the Ages and Stages Questionnaire (third edition) to the VDAART primary caregivers during a study clinic visit or phone interview. The ASQ assesses 5 developmental domains (gross motor skills, fine motor skills, problem-solving ability, communication, and personal and social skills) at different ages (year one, n=99; year two, n=102; and year three, n=110). Each domain is assessed by 6 questions ascertaining achievement of relevant skills and answered as yes (10 points), sometimes (5 points), or not yet (0 points). Scores for individual items (within the same domains) are summed to give an overall continuous score for each of the 5 domains (possible range, 0-60).

### Sequencing and profiling of Bacterial 16S rRNA

Metagenomic sequencing and profiling was performed on stool samples by sequencing the 16S rRNA hypervariable region 4 (V4) on the Illumina MiSeq platform, as previously described^42^. Raw reads were demultiplexed and processed (e.g., quality filtering and trimming) using tools available in QIIME 2 (version 2020.8)^43^. Clean reads (averagely 51,935 ± 24,962 range from 23,983 to 179,820) were then denoised and clustered with dada2 and classified using a pre-trained machine-learning-based classifier of the Silva database.

### Measurement of Short Chain Fatty Acids (SCFA)

Eight SCFA were measured by liquid chromatography with tandem mass spectrometry by Metabolon, Inc (Morrisville, NC): acetic acid, propionic acid, butyric acid, isobutyric acid, 2-methyl-butyric acid, isovaleric acid, valeric acid, and hexanoic acid. Sample preparation was conducted according to previously described methods^44^. In brief, the reaction mixture was diluted, and an aliquot injected onto an Agilent 1290/AB Sciex QTrap 5500 liquid chromatography with tandem mass spectrometry system equipped with a C18 reversed phase ultra-high performance liquid chromatography column. Raw data were collected and processed using AB SCIEX software Analyst 1.6.2.

### Metabolomics profiling

Stool samples taken from the selected paired mothers and children (**Fig.1**) were used for metabolomic profiling. Sample preparation was conducted according to previously described methods^45^. Nontargeted global metabolomic profiles were generated at Metabolon Inc. by using ultra-performance liquid chromatography–tandem mass spectroscopy (UPLC-MS/MS). Briefly, four platforms (UPLC-MS/MS under positive ionization, UPLC-MS/MS under negative ionization, UPLC-MS/MS polar platform, and gas chromatography-MS) were used to detect a comprehensive list of metabolites throughout the metabolome. Metabolites were identified by their m/z, retention time, and through a comparison to library entries of purified known standards. For the data preparation and quality control, metabolites with CV >25%, missingness >10%, or no variability based on the IQR we removed in the quality control samples. Metabolon’s authentic standard library contains identifying ion and chromatographic features of over 4,000 known compounds (and an additional 7,000 entries for unnamed compounds) present in metabolic pathways.

### Statistical analysis

The Bray-Curtis dissimilarity matrices (based on ASVs, metabolites, and SCFAs) were calculated by QIIME2. PERMANOVA (Adonis) was then used to test the effect size of factors such as recruitment site, education level, marital status, family income, gestational age, Vitamin D treatment, mothers’ health status and children’s ASQ measures on the multi-omics data. We chose not to use classic multiple comparison correction techniques (e.g., Bonferroni) because these would be too conservative, given the correlated nature of the outcomes and the compositionality of microbiome and metabolome data, and because these analyses were performed for the purposes of covariate selection and not for causal inference.

### Multinomial regression

Songbird^26^ (https://github.com/biocore/songbird) was used to perform the multinomial regression including null model construction. We included as covariates all subject characteristics that were associated with gut microbiota and metabolites (PERMANOVA test P-value < 0.05 or F value > 2, **Table S1**). The model with these covariates was compared with a null model containing no covariates to estimate a Q^2^ adapted from the partial least squares literature. Specifically, Q^2^ = 1 - m1/m2, where m1 indicates the average absolute model error and m2 indicates the average absolute null model error. Q^2^ values close to 1 indicate a high predictive accuracy on cross-validation samples. Q^2^ values that are low or below zero indicate poor predictive accuracy, suggesting possible overfitting.

The output from Songbird also includes differentials which describe the log-fold change of microbes/metabolites with respect to ASQ measures. The most important aspect of these differentials are ranks (Songbird does not generate p values but instead generates taxa’s or metabolites’ ranks because when using a reference-frame based approach, there is no reasonable null distribution for generation of test statistics), which are obtained by sorting a column of differentials from lowest to highest. These ranks give information on the relative associations of features (microbes and metabolites) with a given covariate e.g., the features with the most negative differential ranking values will be more associated with low ASQ samples, whereas the features with the most positive differential ranking values will be more associated with high ASQ samples.

## Data Availability

All data produced in the present study are available upon reasonable request to the authors.

## Notes

### Competing Interest Statement

The authors have declared no competing interest.

### Funding Statement

This study was funded by grants from the National Institutes of Health (R01AI141529, R01HD093761, RF1AG067744, UH3OD023268, U19AI095219, and U01HL089856) and the Charles A. King Trust Postdoctoral Fellowship.

### Author Declarations

Subjects were offspring of participants in VDAART, a multi-site randomized, double-blind, placebo-controlled trial of Vitamin D supplementation during pregnancy for prevention of asthma and other allergic diseases in offspring conducted in the United States (NCT00920621). The study protocol was approved by the institutional review boards at each participating institution and at Brigham and Women's Hospital (from three sites across the United States: Boston Medical Center, Washington University in St. Louis, and Kaiser Permanente Southern California Region in San Diego). All participants provided written informed consent.

## Reference

1. Clemente, J.C., Ursell, L.K., Parfrey, L.W. & Knight, R. The Impact of the Gut Microbiota on Human Health: An Integrative View. Cell 148, 1258–1270 (2012).

2. Sherwin, E., Bordenstein, S.R., Quinn, J.L., Dinan, T.G. & Cryan, J.F. Microbiota and the social brain. Science (New York, N.Y.) 366(2019).

3. Collins, S.M., Surette, M. & Bercik, P. The interplay between the intestinal microbiota and the brain. Nature reviews. Microbiology 10, 735–742 (2012).

4. Braniste, V., et al. The gut microbiota influences blood-brain barrier permeability in mice. Science translational medicine 6, 263ra158 (2014).

5. Chu, C., et al. The microbiota regulate neuronal function and fear extinction learning. Nature 574, 543-+ (2019).

6. Erny, D., et al. Host microbiota constantly control maturation and function of microglia in the CNS. Nat Neurosci 18, 965-+ (2015).

7. Hoban, A.E., et al. Regulation of prefrontal cortex myelination by the microbiota. Transl Psychiat 6(2016).

8. Ogbonnaya, E.S., et al. Adult Hippocampal Neurogenesis Is Regulated by the Microbiome. Biol Psychiat 78, E7–E9 (2015).

9. Cryan, J.F. & Dinan, T.G. Mind-altering microorganisms: the impact of the gut microbiota on brain and behaviour. Nat Rev Neurosci 13, 701–712 (2012).

10. Clarke, G., et al. The microbiome-gut-brain axis during early life regulates the hippocampal serotonergic system in a sex-dependent manner. Mol Psychiatr 18, 666–673 (2013).

11. Sudo, N., Chida, Y. & Kubo, C. Postnatal microbial colonization programs the hypothalamic-pituitary-adrenal system for stress response in mice. J Psychosom Res 58, S60–S60 (2005).

12. Heijtz, R.D. Fetal, neonatal, and infant microbiome: perturbations and subsequent effects on brain development and behavior. Semin Fetal Neonat M 21, 410–417 (2016).

13. Buffington, S.A., et al. Microbial Reconstitution Reverses Maternal Diet-Induced Social and Synaptic Deficits in Offspring. Cell 165, 1762–1775 (2016).

14. Yu, L., Zhong, X., He, Y. & Shi, Y. Butyrate, but not propionate, reverses maternal diet-induced neurocognitive deficits in offspring. Pharmacological research 160, 105082 (2020).

15. Kim, S., et al. Maternal gut bacteria promote neurodevelopmental abnormalities in mouse offspring. Nature 549, 528-+ (2017).

16. Dawson, S.L., et al. Maternal prenatal gut microbiota composition predicts child behaviour. EBioMedicine 68, 103400 (2021).

17. Meckel, K.R. & Kiraly, D.D. Maternal microbes support fetal brain wiring. Nature 586, 203–205 (2020).

18. Jasarevic, E., et al. The maternal vaginal microbiome partially mediates the effects of prenatal stress on offspring gut and hypothalamus. Nat Neurosci 21, 1061-+ (2018).

19. Tochitani, S. Vertical transmission of gut microbiota: Points of action of environmental factors influencing brain development. Neurosci Res 168, 83–94 (2021).

20. Vuong, H.E., et al. The maternal microbiome modulates fetal neurodevelopment in mice. Nature 586, 281-+ (2020).

21. Luczynski, P., et al. Growing up in a Bubble: Using Germ-Free Animals to Assess the Influence of the Gut Microbiota on Brain and Behavior. Int J Neuropsychoph 19(2016).

22. Walter, J., Armet, A.M., Finlay, B.B. & Shanahan, F. Establishing or Exaggerating Causality for the Gut Microbiome: Lessons from Human Microbiota-Associated Rodents. Cell 180, 221–232 (2020).

23. Litonjua, A.A., et al. The Vitamin D Antenatal Asthma Reduction Trial (VDAART): Rationale, design, and methods of a randomized, controlled trial of vitamin D supplementation in pregnancy for the primary prevention of asthma and allergies in children. Contemporary clinical trials 38, 37–50 (2014).

24. Granana, N. & Romero Otalvaro, A.M. Chapter 28 - Neurodevelopment and the Ages and Stages Questionnaire, third edition (ASQ-3). in Diagnosis, Management and Modeling of Neurodevelopmental Disorders (eds. Martin, C.R., Preedy, V.R. & Rajendram, R.) 319–328 (Academic Press, 2021).

25. Landa, R.J., Gross, A.L., Stuart, E.A. & Bauman, M. Latent class analysis of early developmental trajectory in baby siblings of children with autism. J Child Psychol Psyc 53, 986–996 (2012).

26. Morton, J.T., et al. Establishing microbial composition measurement standards with reference frames. Nat Commun 10, 2719 (2019).

27. Mandal, S., et al. Analysis of composition of microbiomes: a novel method for studying microbial composition. Microb Ecol Health Dis 26, 27663 (2015).

28. Morton, J.T., et al. Balance Trees Reveal Microbial Niche Differentiation. mSystems 2(2017).

29. Silverman, J.D., Durand, H.K., Bloom, R.J., Mukherjee, S. & David, L.A. Dynamic linear models guide design and analysis of microbiota studies within artificial human guts. Microbiome 6, 202 (2018).

30. Aijo, T., Muller, C.L. & Bonneau, R. Temporal probabilistic modeling of bacterial compositions derived from 16S rRNA sequencing. Bioinformatics 34, 372–380 (2018).

31. Xia, F., Chen, J., Fung, W.K. & Li, H. A logistic normal multinomial regression model for microbiome compositional data analysis. Biometrics 69, 1053–1063 (2013).

32. Cheung, S.G., et al. Systematic Review of Gut Microbiota and Major Depression. Front Psychiatry 10, 34 (2019).

33. Katz Sand, I., et al. Disease-modifying therapies alter gut microbial composition in MS. Neurol Neuroimmunol Neuroinflamm 6, e517 (2019).

34. El Aidy, S., et al. Serotonin Transporter Genotype Modulates the Gut Microbiota Composition in Young Rats, an Effect Augmented by Early Life Stress. Front Cell Neurosci 11, 222 (2017).

35. Hughes, H.K., Rose, D. & Ashwood, P. The Gut Microbiota and Dysbiosis in Autism Spectrum Disorders. Curr Neurol Neurosci Rep 18, 81 (2018).

36. Bezawada, N., Phang, T.H., Hold, G.L. & Hansen, R. Autism Spectrum Disorder and the Gut Microbiota in Children: A Systematic Review. Ann Nutr Metab 76, 16–29 (2020).

37. Zhang, W., et al. Preliminary evidence for an influence of exposure to polycyclic aromatic hydrocarbons on the composition of the gut microbiota and neurodevelopment in three-year-old healthy children. BMC Pediatr 21, 86 (2021).

38. Hsiao, E.Y., et al. Microbiota modulate behavioral and physiological abnormalities associated with neurodevelopmental disorders. Cell 155, 1451–1463 (2013).

39. Kelly, J.R., Minuto, C., Cryan, J.F., Clarke, G. & Dinan, T.G. Cross Talk: The Microbiota and Neurodevelopmental Disorders. Front Neurosci 11, 490 (2017).

40. Ding, X., et al. Gut microbiota changes in patients with autism spectrum disorders. J Psychiatr Res 129, 149–159 (2020).

41. Qiao, Y., et al. Effect of combined chronic predictable and unpredictable stress on depression-like symptoms in mice. Ann Transl Med 8, 942 (2020).

42. Lee-Sarwar, K.A., et al. Integrative analysis of the intestinal metabolome of childhood asthma. J Allergy Clin Immunol 144, 442–454 (2019).

43. Bolyen, E., et al. Reproducible, interactive, scalable and extensible microbiome data science using QIIME 2. Nat Biotechnol 37, 852–857 (2019).

44. Lee-Sarwar, K.A., et al. Fecal short-chain fatty acids in pregnancy and offspring asthma and allergic outcomes. J Allergy Clin Immunol Pract 8, 1100–1102 e1113 (2020).

45. Kelly, R.S., et al. Asthma Metabolomics and the Potential for Integrative Omics in Research and the Clinic. Chest 151, 262–277 (2017).

