## Supplemental materials for "Identifying the critical time window for the association of the early-life gut microbiome and metabolome with childhood neurodevelopment"

**Supplemental Tables and Figures**

**Figure S1.** Heatmap depicting availability of multi-omics data of stool samples and ASQ questionnaires for 116 paired mothers and children.

**Figure S2.** The scatter plots of PCoA for the gut microbiota and metabolites based on the Bray-Curtis dissimilarity.

**Figure S3.** Details of predictive power (Q2) of the gut microbiome at different taxonomy levels for neurodevelopment measured by ASQ.

**Table S1.** The F values of the Permanova Test for all covariates in each sub-datasets for multinomial regression.

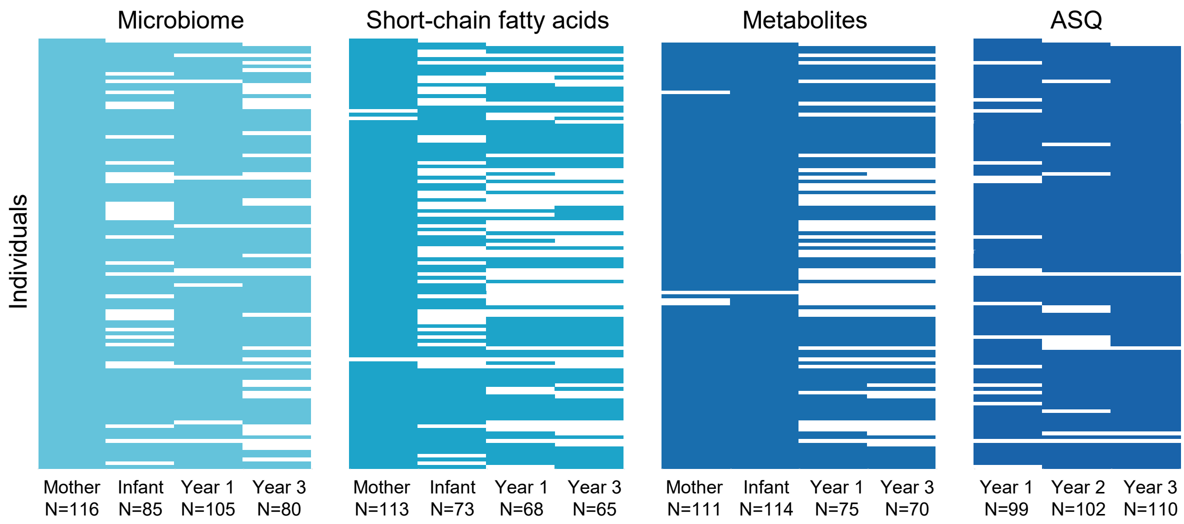


**Figure S1. Heatmap depicting availability of multi-omics data of stool samples and ASQ questionnaires for 116 paired mothers and children.** The X-axis refers to different sampling stages, and the Y-axis refers to paired mothers and children. From left to right, blue blocks indicate available multi-omics data of stool samples and the ASQ measures, respectively, while white represents missing data.

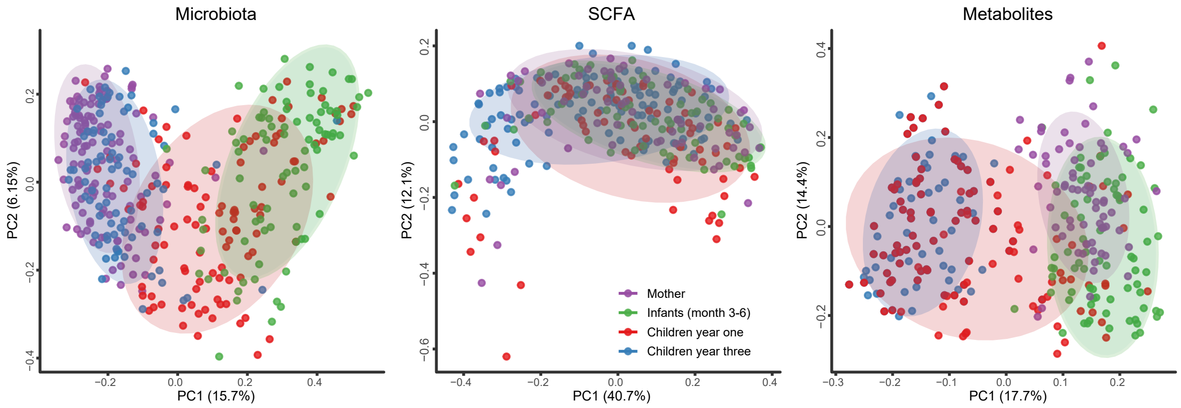


**Figure S2. The scatter plots of PCoA for the gut microbiota and metabolites based on the Bray-Curtis dissimilarity.** F value and *p* value of the Permanova test are marked on the plots as well to quantify the differences of metagenomics and metabolomics data among different stages (purple dots for maternal samples, green for infant, red for children at year one, and blue for children at year three).

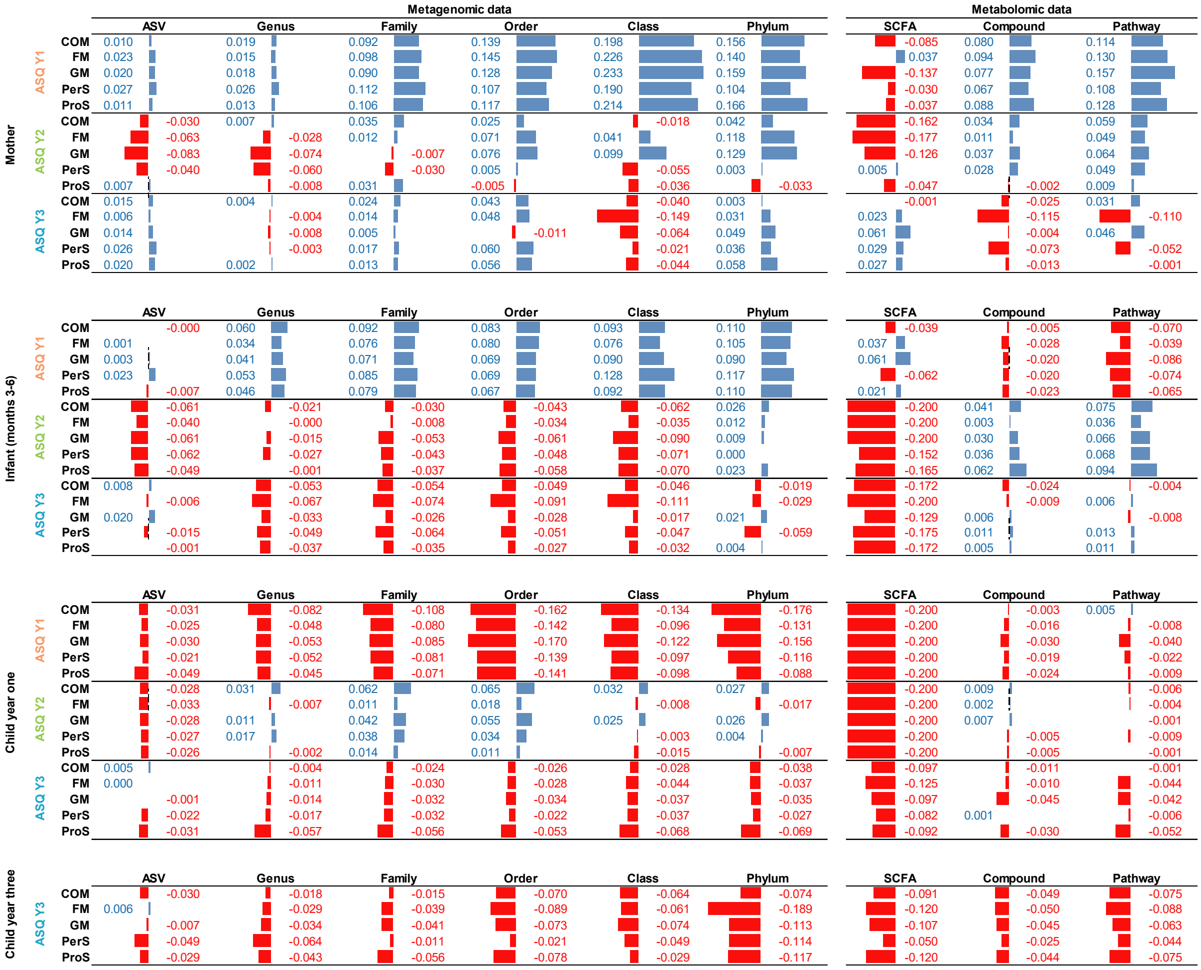


**Figure S3. Details of predictive power ($Q^2$) of the gut microbiome at different taxonomy levels for neurodevelopment measured by ASQ.** (**a**) Predictive power of the maternal gut microbiome for child neurodevelopment. (**b**) Predictive power of the infant gut microbiome in early life (months 3-6) for child neurodevelopment. (**c**) Predictive power of the child gut microbiome at year one for child neurodevelopment. (**d**) Predictive power of the child gut microbiome at year three for child neurodevelopment at the same time point. Red font and bar indicate negative $Q^2$ and green indicates positive $Q^2$. COM, FM, GM, PerS, and ProS are short for communication skills, fine motor skills, gross motor skills, personal social skills, and problem-solving skills, respectively. The higher $Q^2$ (positive) indicates a higher predictive accuracy for ASQ, while negative $Q^2$ indicates poor predictive accuracy or overfitting of the multinomial regression models for ASQ measures. The value less than -0.2 will be trimmed to -0.2 for visualization.

| | | Microbiota | | | | | | Scfas | Metabolites | |
|---|---|---|---|---|---|---|---|---|---|---|
| | | ASV | Genus | Family | Order | Class | Phylum | SCFAs | Comps | Pathways |
| Mother | Age | 2.046 | 2.413 | 2.831 | 3.058 | 3.083 | 2.818 | 1.052 | 3.502 | 3.591 |
| | Recruitment site | 1.685 | 1.612 | 1.477 | 1.596 | 1.551 | 1.781 | 2.569 | 1.485 | 1.425 |
| | Education level | 1.309 | 1.497 | 1.533 | 1.429 | 1.495 | 1.471 | 0.738 | 1.695 | 1.615 |
| | Marital status | 1.543 | 1.734 | 1.583 | 1.950 | 2.315 | 2.628 | 0.635 | 1.655 | 1.577 |
| | Family income | 1.136 | 1.161 | 1.196 | 1.139 | 1.078 | 1.147 | 1.116 | 1.721 | 1.838 |
| | Vitamin D treatment | 0.970 | 0.879 | 0.542 | 0.662 | 0.221 | -0.017 | 0.257 | 0.801 | 0.888 |
| | Ethnicity | 1.538 | 1.563 | 1.403 | 1.384 | 1.149 | 1.938 | 0.249 | 3.731 | 4.312 |
| | Race | 1.384 | 1.582 | 1.461 | 1.795 | 2.164 | 2.306 | 0.903 | 1.524 | 1.447 |
| | Asthma | 0.928 | 0.912 | 1.246 | 1.438 | 1.514 | 1.795 | 0.285 | 0.598 | 0.533 |
| | Fever | 0.693 | 0.766 | 0.735 | 0.714 | 0.737 | 1.033 | 0.795 | 0.807 | 0.840 |
| | Eczema | 1.109 | 1.185 | 1.423 | 1.357 | 1.446 | 1.279 | 0.670 | 0.692 | 0.697 |
| | Gestation days | 0.883 | 0.791 | 0.866 | 0.338 | 0.748 | 0.739 | 1.335 | 0.594 | 0.498 |
| | Delivery mode | 0.757 | 0.675 | 0.629 | 0.632 | 0.709 | 0.968 | 0.538 | 0.760 | 0.730 |
| | Breast feed | 1.448 | 1.945 | 1.672 | 1.605 | 2.251 | 2.216 | 0.368 | 1.407 | 1.135 |
| | Ethnicity (child) | 1.551 | 1.950 | 2.025 | 2.352 | 2.617 | 4.028 | 0.367 | 3.977 | 4.609 |
| | Race (child) | 1.534 | 1.720 | 1.582 | 1.919 | 2.389 | 2.609 | 0.592 | 1.666 | 1.620 |
| | Gender (child) | 1.636 | 1.347 | 1.330 | 1.484 | 2.308 | 1.199 | 0.796 | 1.305 | 1.311 |
| Infant (months 3-6) | Age | 1.648 | 3.054 | 3.652 | 4.685 | 3.962 | 4.846 | 6.358 | 1.842 | 2.072 |
| | Recruitment site | 1.606 | 1.633 | 1.702 | 1.615 | 1.720 | 1.693 | 1.664 | 1.021 | 1.124 |
| | Education level | 1.208 | 1.191 | 1.298 | 1.486 | 1.293 | 1.500 | 1.960 | 1.221 | 1.215 |
| | Marital status | 1.584 | 1.072 | 1.132 | 1.176 | 1.152 | 0.512 | 3.364 | 1.826 | 2.434 |
| | Family income | 1.091 | 1.004 | 1.179 | 1.225 | 1.148 | 0.904 | 2.493 | 1.478 | 1.640 |
| | Vitamin D treatment | 0.877 | 0.786 | 0.772 | 0.755 | 0.728 | 0.975 | 0.179 | 1.360 | 1.765 |
| | Ethnicity | 1.667 | 1.397 | 1.301 | 1.452 | 1.412 | 2.165 | 0.277 | 2.348 | 2.281 |
| | Race | 1.643 | 1.236 | 1.153 | 1.133 | 1.137 | 0.967 | 1.439 | 1.227 | 1.415 |
| | Asthma | 0.806 | 0.657 | 0.523 | 0.796 | 0.676 | 0.897 | 1.584 | 1.027 | 0.928 |
| | Fever | 1.793 | 1.709 | 0.711 | 0.713 | 0.661 | 1.057 | 3.279 | 1.943 | 1.894 |
| | Eczema | 1.069 | 1.500 | 1.739 | 1.898 | 1.855 | 1.676 | 1.360 | 1.085 | 0.914 |
| | Gestation days | 1.151 | 1.696 | 2.131 | 2.043 | 2.007 | 2.969 | 0.104 | 1.391 | 1.541 |
| | Delivery mode | 1.347 | 1.601 | 1.266 | 1.579 | 1.317 | 1.386 | 0.671 | 0.595 | 0.789 |
| | Breast feed | 2.637 | 2.664 | 3.553 | 4.272 | 3.662 | 3.978 | 6.491 | 2.079 | 2.317 |
| | Ethnicity (child) | 1.731 | 1.471 | 1.621 | 1.719 | 1.752 | 2.730 | 0.579 | 2.230 | 2.284 |
| | Race (child) | 1.588 | 1.265 | 1.333 | 1.315 | 1.307 | 1.103 | 1.998 | 1.711 | 1.833 |
| | Gender (child) | 0.756 | 0.939 | 1.084 | 1.363 | 1.186 | 0.724 | 0.386 | 1.789 | 1.823 |
| Child year one | Age | 1.539 | 1.566 | 1.423 | 1.337 | 1.257 | 0.676 | 1.266 | 1.268 | 1.111 |
| | Recruitment site | 1.498 | 1.470 | 1.764 | 1.509 | 1.021 | 0.136 | 2.304 | 1.046 | 0.821 |
| | Education level | 1.163 | 1.320 | 1.562 | 1.472 | 1.194 | 0.738 | 0.937 | 0.904 | 0.874 |
| | Marital status | 1.611 | 1.566 | 1.481 | 1.455 | 1.312 | 0.910 | 5.613 | 1.231 | 1.127 |
| | Family income | 1.227 | 1.261 | 1.399 | 1.405 | 1.344 | 1.328 | 3.305 | 1.097 | 1.129 |
| | Vitamin D treatment | 0.593 | 0.661 | 0.990 | 1.020 | 0.699 | 0.965 | 0.583 | 1.531 | 1.650 |
| | Ethnicity | 0.792 | 0.641 | 0.475 | 0.565 | 0.947 | 0.995 | 0.367 | 0.482 | 0.309 |
| | Race | 1.267 | 1.338 | 0.992 | 0.971 | 0.680 | 0.465 | 2.140 | 0.906 | 0.716 |
| | Asthma | 0.646 | 0.466 | 0.559 | 0.622 | 0.827 | 0.366 | 1.188 | 0.811 | 0.721 |
| | Fever | 0.919 | 0.814 | 0.844 | 0.649 | 0.871 | 0.430 | 3.715 | 0.578 | 0.611 |
| | Eczema | 1.086 | 1.149 | 1.464 | 1.335 | 1.499 | 1.107 | 1.320 | 0.630 | 0.802 |
| | Gestation days | 0.993 | 0.929 | 0.757 | 0.602 | 0.534 | 0.493 | 0.027 | 1.894 | 2.234 |
| | Delivery mode | 2.112 | 3.115 | 3.634 | 4.320 | 3.798 | 2.836 | 0.392 | 0.954 | 0.813 |
| | Breast feed | 2.256 | 2.117 | 2.179 | 2.127 | 2.010 | 0.335 | 3.339 | 1.870 | 1.492 |
| | Ethnicity (child) | 1.059 | 0.840 | 0.540 | 0.592 | 0.886 | 0.848 | 0.427 | 0.473 | 0.286 |
| | Race (child) | 1.350 | 1.652 | 1.385 | 1.339 | 1.081 | 0.859 | 2.763 | 1.289 | 1.155 |
| | Gender (child) | 0.968 | 0.836 | 0.892 | 0.636 | 0.554 | 0.483 | 1.456 | 0.510 | 0.368 |
| Child year three | Age | 1.206 | 0.813 | 0.694 | 0.428 | 0.054 | 0.048 | 0.457 | 1.581 | 1.504 |
| | Recruitment site | 1.540 | 1.910 | 1.915 | 2.006 | 2.164 | 2.334 | 0.854 | 1.502 | 1.417 |
| | Education level | 1.168 | 1.054 | 0.851 | 0.647 | 0.666 | 0.598 | 0.848 | 1.377 | 1.469 |
| | Marital status | 1.220 | 1.113 | 1.059 | 0.831 | 1.187 | 1.269 | 1.459 | 1.603 | 1.645 |
| | Family income | 1.026 | 0.913 | 0.639 | 0.518 | 0.424 | 0.333 | 1.029 | 1.607 | 1.769 |
| | Vitamin D treatment | 0.780 | 0.655 | 0.397 | 0.305 | 0.511 | 0.975 | 0.304 | 1.275 | 1.283 |
| | Ethnicity | 0.737 | 0.794 | 0.641 | 0.342 | 0.525 | 0.583 | 0.236 | 1.183 | 0.973 |
| | Race | 1.226 | 1.145 | 0.984 | 0.953 | 0.977 | 1.008 | 1.116 | 1.532 | 1.585 |
| | Asthma | 0.811 | 0.531 | 0.453 | 0.445 | 0.615 | 0.837 | 0.131 | 0.971 | 0.963 |
| | Fever | 0.817 | 0.784 | 1.001 | 1.153 | 1.473 | 1.415 | 0.713 | 1.226 | 0.843 |
| | Eczema | 0.822 | 0.984 | 0.668 | 0.642 | 0.588 | 0.239 | 0.428 | 0.827 | 0.779 |
| | Gestation days | 1.446 | 2.098 | 2.452 | 3.044 | 2.978 | 3.500 | 0.517 | 0.982 | 0.746 |
| | Delivery mode | 0.980 | 0.745 | 0.775 | 0.300 | 0.484 | 0.327 | 3.211 | 1.512 | 1.428 |
| | Breast feed | 1.545 | 1.537 | 1.191 | 0.588 | 0.468 | 0.564 | 1.097 | 2.353 | 2.261 |
| | Ethnicity (child) | 0.769 | 0.761 | 0.650 | 0.352 | 0.430 | 0.458 | 0.245 | 1.376 | 1.131 |
| | Race (child) | 1.234 | 1.080 | 0.878 | 0.738 | 0.881 | 0.803 | 0.333 | 1.537 | 1.689 |
| | Gender (child) | 1.073 | 1.281 | 1.519 | 1.518 | 0.900 | 0.926 | 3.817 | 1.090 | 0.937 |

**Table S1. The F values of the Permanova Test for all covariates in each sub-datasets for multinomial regression.**
